## Supplementary material for "A Stakeholder-Led Understanding of the Implementation of Digital Technologies Within Heart Disease Diagnosis: A Qualitative Study Protocol": (see Appendix)

Digital Twin Question Schedules

Lived Experience Focus Group Questions

Heart Disease-related Questions (diagnosis process)

1. Please describe the series of events which took place that led to your heart disease diagnosis
   - Possible prompts: how involved did you feel in this process, active/passive, taken seriously by clinicians/dismissed, big/small role

2. How did you keep track of your symptoms before you were given your diagnosis?
   - Possible prompts: digital tools used, manual log, own method

3. After you first brought up concerns about your heart to your doctor, what did they ask you to do, if anything?
   - Possible prompts: digital tools used, manual log, own method

4. What types of numeric data did your doctor discuss with you, if any?
   - Possible prompts: percentage risks, heart rates, blood pressure, etc
   - Follow up: Was the meaning/importance of the data clear to you?

5. What other aspects of your overall health and wellbeing were important to you around the time you were getting diagnosed?
   - Possible prompts: existing comorbidities, mental health, social support, stress, financial status, lifestyle choices, health behaviours etc

6. During the diagnosis process, what were some important things/people that made the experience less difficult?
   - Possible prompts: staff, communication, social support at home

7. What were the greatest challenges for you during the diagnosis process?
   - Possible prompts: communication, access to information/emotional support, dealing with clinicians, difficulty understanding medical terminology

8. What were your experiences with doctors, nurses and other healthcare staff during the process of getting diagnosed?
   - Possible prompts: positive/negative experience, specific experiences standing out, communication, prejudices or discrimination?
   - Follow up (depending on previous responses) – how did this impact the rest of your treatment?
   - Follow up (depending on previous responses) – how has communication with your clinician impacted your treatment?

9. What do you feel delays or prevents people from getting accurate diagnoses?
   - Possible prompts: access to healthcare, language barriers, fear of stigma/discrimination, lack of concern for health, financial burdens, lack of social support

10. How do you think we could address these things to help someone get a quicker and more accurate diagnosis?
    - Possible prompts: better, more objective data, easier to explain problems to clinicians, access to healthcare, language assistant, addressing stigmas, financial support, social/mental health/emotional support

Digital Technologies Questions

1. Technologies such as televisions, computers, smartphones etc. have become increasingly integral to our daily lives.
   How do you feel about using different types of technologies in your life?
    - Possible prompts: positive/negative, certain elements easier/harder

2. More specifically, digital technologies are those that can electronically generate, process and store different types of data. Digital technologies are increasingly being used in the healthcare sector.
   What experiences do you have using digital health tools within your healthcare treatment (doesn't have to be specific to heart disease diagnosis)?
    - Possible prompts: positive/negative, limited use, fit for purpose, implementation by clinicians, clinician experience with them

3. Thinking back to the challenges we mentioned in the first part of our discussions [give examples], do you feel digital technologies would help improve, or make worse, any of these issues? If so, which ones and how?
    - Note down issues mentioned and prompt those if needed

Digital health tools are being used more and more in the healthcare sector, as they can improve the way we diagnose and monitor many conditions (including heart disease) by tracking symptoms more effectively.

4. If you were given a digital health tool to help improve the accuracy of your heart disease diagnosis, what sort of things would put you off from using it?
    - Possible prompts: digital literacy, not seen as valuable, preference to deal with a real person, health status, financial status

5. What would motivate you to use it?
    - Possible prompts: incentives, feedback, avatar

6. How would you feel about getting feedback from this type of technology?
    - Follow up: how would it impact your engagement with it?
    - Possible prompts: agency in diagnosis, better awareness of condition/symptoms, more accurate monitoring

7. How would you feel about getting feedback from this device before seeing your doctor?
    - Possible prompts: examples like not enough movement, try to stand more etc;

8. Who do you think should receive the information that comes from this device?
    - Prompts: clinician, patient, family

9. Do you think there will be any challenges when trying to include a digital monitoring tool in daily patient use?
    - Follow up: What factors do you think we need to consider to make it as easy to use as possible?

The final two questions will be more specific to the final product we are aiming to create. Try to imagine you have a wearable device that is linked to a smartphone app, which allows you to enter data about your symptoms, how you are feeling, certain stressors that may be occurring in your day and so on. This data would all be processed by the app and then reported back to you and your clinician in an easy-to-read format. Let’s discuss what you think about specific elements that might be included…

10. If the data were to be sent to you, what would be the most helpful way for the data to be presented/communicated to you?
    - Possible prompts: features like colours, layout etc., how to communicate it – email summary, app interface etc, would you want to see it yourself before seeing your clinician or not?

11. What are your thoughts on having a virtual doctor/avatar/chatbot to interact with on a device?
    - Follow up: do you think being able to personalise the avatar would help with engagement and relatability of the virtual element?
    - Follow up: do you think this would be better than having a simple logging/tracking system like they use for other health tracking apps? Or more complicated?
    - Follow up: any immediate deterrents come to mind from having a ‘chatbot’ type function?

Digital Twin Question Schedules

Clinician Interview Questions

Heart Disease-related Questions (diagnosis process)

1. Please describe the series of events that normally take place when you diagnose a patient with heart disease.
    - Possible prompts: factors into your decision, length of time, with collaboration/advice from other colleagues, how it's changed with years of experience.

2. How do you monitor your patients' symptoms prior to/following diagnosis?
    - Follow up: what do you ask them to do?
    - Possible prompts: digital tools used, manual log, own method

3. Which numeric data do you find most valuable when making diagnoses?
    - Possible prompts: ECGs, heart rate, blood pressure etc.

4. What other aspects of overall patient health and wellbeing are important to you when you are making a heart disease diagnosis?
    - Possible prompts: existing comorbidities, mental health, social support, stress, financial status, lifestyle choices, health behaviours etc

5. What do you feel are important factors for ensuring a patient has a positive experience following a heart disease diagnosis?
    - Possible prompts: staff, communication, social support at home

6. What are the greatest challenges for you, as a clinician, during the diagnosis process?
    - Possible prompts: communication, medical advice adherence, language barriers, implicit biases due to potentially 'self-inflicted' nature of disease

7. What are your interactions with your patients like prior to/following their heart disease diagnosis?
    - Possible prompts: positive/negative experience, specific experiences standing out, communication, prejudices or discrimination?
    - Follow up (depending on previous responses) – how did this impact the rest of their treatment?
    - Follow up (depending on previous responses) – how has communication with your patients impacted your treatment?

8. What do you feel are potential barriers that prevent people from getting accurate diagnoses?
    - Possible prompts: access to healthcare, language barriers, fear of stigma/discrimination, lack of concern for health, financial burdens, lack of social support

9. How do you think we could address these barriers to help someone get an accurate diagnosis faster?
    - Possible prompts: better, more objective data, easier to explain problems to clinicians, access to healthcare, language assistant, addressing stigmas, financial support, social/mental health/emotional support

Digital Technologies Questions

1. Technologies such as televisions, computers, smartphones etc. have become increasingly integral to our daily lives.
   How do you feel about using different types of technologies in your life?
    - Possible prompts: positive/negative, certain elements easier/harder

2. What experiences do you have using digital health tools within your healthcare profession? (doesn't have to be specific to heart disease diagnosis)
    - Possible prompts: positive/negative, limited use, fit for purpose, implementation by clinicians, clinician experience with them

3. Thinking back to the challenges we mentioned in the first part of our discussions [give examples], do you feel digital technologies would help improve, or make worse, any of these issues? If so, which ones and how?
    - Note down issues mentioned and prompt those if needed

As you may know, digital health tools are being used more and more in the healthcare sector, to improve the way we diagnose and monitor many conditions (including heart disease) by tracking symptoms more effectively.

4. What would prevent you from using/prescribing a digital health tool made for improving diagnosis of heart disease?
    - Possible prompts: digital literacy, not seen as valuable, preference to deal with a real person, health status, financial status

5. What do you think would prevent your patients from engaging with a digital health tool for symptom monitoring?
    - Possible prompts: digital literacy, not seen as valuable, preference to deal with a real person, health status, financial status

6. What would motivate you to engage in such technologies?
    - Possible prompts: incentives, feedback, avatar, certain data types/presentation/summaries

7. What do you think would motivate your patients to engage in such technologies?
    - Possible prompts: incentives, feedback, avatar

8. Do you feel that patients getting feedback from a digital monitoring system would improve engagement with it? If so, how, why/why not?
    - Possible prompts: agency in diagnosis, better awareness of condition/symptoms, more accurate monitoring

9. How would you feel if your patients were getting feedback from the digital monitoring system before seeing you?
    - Possible prompts: examples like not enough movement, try to stand more etc;

10. Do you foresee any challenges with implementing a digital monitoring tool into daily patient use? What factors do you think we need to consider to make it as easy to use as possible?

The final two questions will be more specific to the final product we are aiming to create. Try to imagine you will be giving patients a wearable device that is linked to a smartphone app, which allows them to enter data about their symptoms, how they are feeling, certain stressors that may be occurring in their day and so on. This data would all be processed by the app and then reported back to you and your patient in an easy-to-read format. Let's discuss what you think about specific elements that might be included…

11. What are your thoughts on having a virtual doctor/avatar/chatbot to interact with on a device?
    - Follow up: do you think being able to personalise the avatar would help with engagement and relatability of the virtual element?
    - Follow up: do you think this would be better than having a simple logging/tracking system like they use for other health tracking apps? Or more complicated?
    - Follow up: any immediate deterrents come to mind from having a 'chatbot' type function?

12. What would be the most helpful way for the data to be presented/communicated to you?
    - Follow up: how do you think this should differ from how data is presented to patients, if at all?
    - Possible prompts: features like colours, layout etc., how to communicate it – email summary, app interface etc, would you want to see it yourself before seeing your clinician or not?
